# Fragile X Syndrome in Brazil: Development and Validation of a Clinical Checklist for Population Screening

**DOI:** 10.1101/2025.10.21.25338500

**Authors:** Luz María Romero, Gabriela Cristina Carvalho Santos, Vanessa Schubert, Edson Emílio Scalabrin, Roberto Hirochi Herai

## Abstract

Fragile X Syndrome (FXS) is the most common inherited cause of intellectual disability and syndromic autism, but diagnosis remains challenging due to phenotypic variability and limited access to molecular testing in low- and middle-income countries. We conducted a cohort study of 419 individuals with molecularly confirmed full mutation or mosaicism (364 males, 55 females) from the Buko Kaesemodel Institute database to characterize clinical features and develop a screening tool for referral to genetic testing. Twelve physical, cognitive, and behavioral symptoms were assessed, and statistical analyses (95% CI, Pearson correlation) combined with machine learning (Random Forest, K-means) identified the most discriminative traits. The most frequent symptoms were learning difficulties (86.9%), attention deficit (79.7%), delayed speech (73.3%), intellectual disability (72.8%), repetitive movements (70.2%), and characteristic facial features (65.4%). Macroorchidism was present in 41.8% of males. Symptom prevalence in females was similar but with fewer physical features. Moderate correlations were observed, including between intellectual disability and learning difficulties (r = 0.59). A scored checklist, incorporating sex-specific weights, achieved 95% sensitivity with ROC areas of 0.73 (males) and 0.76 (females). Key predictors differed by sex, with intellectual disability and facial features being most informative in males, and learning difficulties in females. This is the largest FXS cohort analyzed in Brazil and provides the most comprehensive symptom profile in an admixed population. The validated checklist represents a practical, low-cost tool to support early diagnosis and guide public health strategies, reducing unnecessary molecular testing while prioritizing high-risk cases.

## INTRODUCTION

Fragile X Syndrome (FXS) (OMIM #300624) is a rare genetic condition and is considered the leading inherited cause of intellectual disability (ID) and syndromic autism (Hagerman & Hagerman, 2002). The etiology of the syndrome is related to expansion of CGG (cytosine–guanine–guanine) trinucleotide repeats in the untranslated 5′UTR region of the FMR1 gene (fragile X messenger ribonucleoprotein 1), located at Xq27.3. This expansion compromises production of the fragile X messenger ribonucleoprotein (FMRP), which is essential for messenger RNA transport, regulation of synaptic plasticity, and maintenance of neuronal function, directly impacting central nervous system development (Deng & Klyachko, 2022; Polussa et al., 2014; Godler & Brown, 2023).

Classification of FMR1 gene alleles varies according to the number of CGG repeats. Individuals with 5 to 44 repeats are considered within the normal range, typically showing adequate FMRP expression and no clinical manifestations related to the syndrome. Those with 45 to 54 repeats are classified in the gray zone and may exhibit subtle changes in gene expression, as well as an increased risk of expansion in subsequent generations (Terraciano et al., 2004; Nolin et al., 2019).

Individuals with 55 to 200 repeats carry a premutation. Although often neurotypical, they may present reduced FMR1 expression and lower FMRP levels, and they are associated with fragile-X–associated primary ovarian insufficiency (FXPOI) and fragile-X–associated tremor/ataxia syndrome (FXTAS), in addition to an elevated risk of expansion to a full mutation in their descendants (Xie et al., 2024). A full mutation occurs when there are more than 200 CGG repeats, generally leading to promoter methylation, silencing of the FMR1 gene, and absence of FMRP. Individuals with a full mutation typically present the FXS clinical phenotype, characterized by a variable combination of cognitive, behavioral, and physical manifestations (Fernandez-Carvajal et al., 2009; Hunter et al., 2023).

It is important to note that the premutation can be transmitted to descendants and may expand to a full mutation (Nolin et al., 2019). Prevalence estimates for the full mutation in the general population, based on studies conducted in Canada and the United States, are approximately 1 in 5,000 boys and between 1 in 4,000 and 1 in 8,000 girls (Hagerman et al., 2017). In Brazil, there are currently no official epidemiological data that precisely describe the occurrence of the condition.

The syndrome manifests heterogeneously across cognitive, physical, and behavioral domains. Frequently observed signs include intellectual disability, hyperactivity with or without attention-deficit symptoms, emotional alterations, and behaviors consistent with autism spectrum disorder (Stone et al., 2019). In addition, affected individuals may exhibit physical features characteristic of the syndrome, notably elongated face, prominent ears, mandibular prognathism, high-arched palate, joint hyperextensibility, and macroorchidism (Hersh & Saul, 2011; Saini et al., 2019; Hagerman & Hagerman, 2021).

Despite descriptions of a “typical” phenotype, it is important to recognize the broad clinical variability of the syndrome. Not all individuals with FXS present the same set of manifestations, which can hinder diagnosis based solely on clinical evaluation (Kanwal et al., 2015; Stone et al., 2023). Given this variability, a definitive diagnosis requires molecular testing. DNA analysis to quantify the number of CGG repeats in the FMR1 gene is fundamental to confirm the causative mutation, especially in cases of intellectual disability without a defined etiology or when there is a family history suggestive of the syndrome across multiple generations (Guruju et al., 2009).

This molecular analysis is performed using polymerase chain reaction (PCR) and/or Southern blotting, from biological samples such as blood, saliva, amniotic fluid, or chorionic villus samples, the latter used in prenatal diagnostic contexts (Jorge, 2013; Hunter et al., 2023). However, these tests are costly and, in developing countries such as Brazil, access to genetic diagnosis remains limited to a small portion of the population. Therefore, the use of screening criteria that enable the prior selection of individuals with a higher probability of harboring the mutation can optimize resources, increase the diagnostic confirmation rate, and expand the provision of appropriate clinical and psychosocial support to patients and their families (Mei et al., 2023).

In this context, since FXS was identified as one of the main causes of intellectual disability, several countries have developed and implemented clinical screening programs based on signs and symptoms suggestive of FXS, with the aim of more accurately indicating which cases should be referred for confirmatory genetic testing. In this study, we conducted a systematic survey of clinical instruments (checklists) used in nine countries (United States, Finland, Belgium, Spain, India, Pakistan, Democratic Republic of the Congo, China, and Brazil), carried out by 12 research groups, to analyze the characteristics reported at higher frequency worldwide.

Hagerman et al. (1991) in the United States evaluated 107 males with intellectual disability or severe learning difficulties and identified 15 cases of Fragile X Syndrome. The most significant findings (p < 0.05) included perseverative speech (93%), large or prominent ears (93%), and macroorchidism (91%). In the same year, Butler et al. (1991), also in the United States, analyzed 188 individuals with ID and identified 19 with a full mutation. Among those with FXS, the most recurrent signs were ID (100%), large ears (94.7%), macroorchidism (84.2%), and hyperactivity (73.7%).

Arvio et al. (1997) in Finland employed a three-step screening protocol (clinical checklist, medical evaluation, and genetic analysis) applied to 197 adults with ID, 26 of whom had FXS, including six newly identified cases. The most frequently observed clinical manifestations were enlarged testes (88%), large ears (77%), and perseverative speech (69%).

Maes et al. (2000) in Belgium validated a checklist composed of 28 items (seven physical and 21 behavioral), using a sample of 110 individuals with FXS and 79 controls, contributing to improved clinical instruments for identifying the syndrome. Lachiewicz et al. (2000) in the United States compared 36 boys with confirmed FXS to 37 children with developmental delay. The most evident signs in the FXS group were soft skin (100%), joint hyperextensibility (100%), and high-arched palate (94%).

Alonso and Gómez (2008) in Spain selected 57 men with ID from an initial sample of 273, identifying 13 FXS-positive cases. Beyond the universal presence of intellectual disability, the most frequent findings were attention-deficit symptoms (96.2%), avoidance of eye contact (85.5%), and perseverative speech (85.5%). In India, Guruju et al. (2009) applied a 15-item clinical checklist to 337 institutionalized individuals with ID, of whom 14 were diagnosed with the syndrome. The main findings included macroorchidism (94.4%), family history of intellectual disability (92%), and enlarged ears (88%).

Kanwal et al. (2015) in Pakistan evaluated 395 individuals with ID and identified 13 with a full mutation of FXS. In addition to the universal presence of ID, hyperactivity (69%), avoidance of eye contact (62%), and speech difficulties (62%) were prominent. In 2018, Lubala et al. conducted a meta-analysis of ten clinical checklists; the most consistent findings across studies included soft skin (88.4%), avoidance of eye contact (86.3%), and large or prominent ears (83.9%). More recently, Mei et al. (2023) in China evaluated 36 children with confirmed FXS. The most frequently reported traits were elongated face (92%), prominent ears (92%), and difficulties in social interaction (75%).

In Brazil, few studies have addressed Fragile X Syndrome using large population samples. Among the most relevant are the works of Mingroni-Netto et al. (1990), Boy et al. (2001), and Christofolini et al. (2007), which contributed to recognition of the syndrome nationally, despite methodological or sampling limitations.

Mingroni-Netto et al. (1990) evaluated 125 students (75 males and 50 females) from schools specialized in serving individuals with intellectual disability. Diagnosis was made by cytogenetic analysis (karyotype), a technique that, although used at the time, has lower sensitivity and specificity compared with current molecular methods. Eight individuals (six males and two females) were diagnosed with FXS, and the study aimed to estimate the frequency of the syndrome among people with ID in Brazil.

Subsequently, Boy et al. (2001) analyzed 104 patients with intellectual disability of idiopathic cause (92 males and 12 females) seen at the Human Genetics Service of the State University of Rio de Janeiro. A full mutation was confirmed in 17 individuals. Among prepubertal boys (n = 4), findings present in all cases included family history of ID, large ears, and avoidance of eye contact. In the post-pubertal group (n = 10), the most frequent manifestations were avoidance of eye contact (100%), elongated face (100%), and family history of ID (90%).

In 2007, Christofolini et al. applied two clinical lists (Butler and Laing) to a sample of 200 men with intellectual disability to evaluate their utility for FXS screening. A full mutation was identified in 19 individuals (9.5%), who had the highest scores on both lists. Among the most recurrent criteria in this group was a family history of intellectual disability, present in 79% of cases.

Despite the advances provided by the checklists developed in the studies presented, these instruments still do not appear to be fully effective for screening. This is due, in part, to the scarcity of clinico-epidemiological investigations that comprehensively address the wide variability of signs and symptoms associated with Fragile X Syndrome. In addition, there is a lack of studies that systematically analyze the manifestations of the syndrome in the Brazilian population.

In this work, we conducted, for the first time in Brazil, a comprehensive survey on FXS with the objective of describing the frequency of the most common signs and symptoms among diagnosed individuals of both sexes. We also performed a combinatorial analysis to identify patterns of co-occurrence and correlations among clinical manifestations. Finally, comparison of the physical, cognitive, and behavioral profiles of males and females enabled the development of a scored checklist with the potential to assist health professionals in clinical recognition of the syndrome and in referral for confirmatory genetic testing.

## METHODS

### Development of the checklist

Based on the evaluation of 12 comparative studies between populations with Fragile X Syndrome (FXS) and control populations (Hagerman, Butler, Arvio, Maes, Lachiewicz, Alonso and Gomez, Guruju, Kanwal, Lubala, Mei, Boy, and Christofolini), we structured the checklist around 12 items most relevant to the Brazilian community, considering physical and behavioral characteristics (Figure 1). A quantitative, descriptive, and comparative analysis was conducted of the most prevalent symptoms in the sample studied.

**Figure 1.**
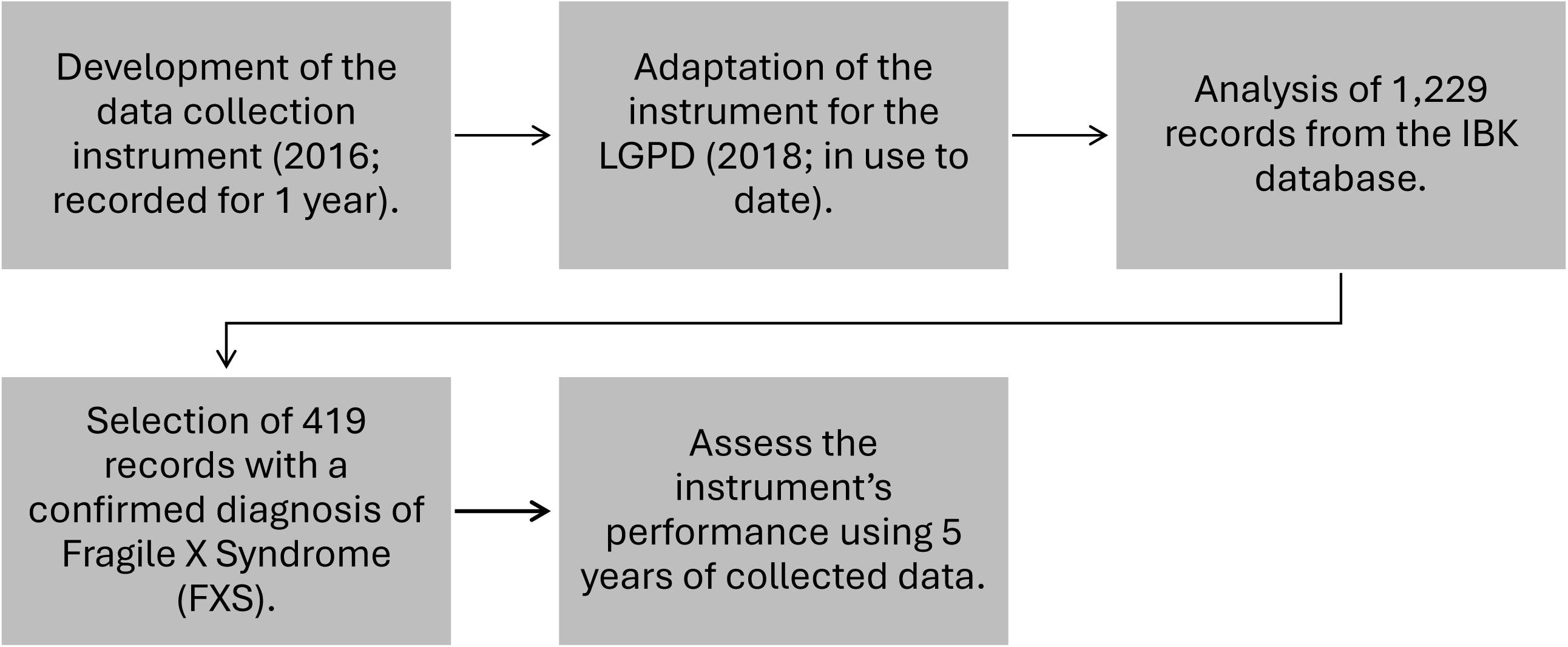
Flowchart of the study data collection strategy.

### Data acquisition

We analyzed records for 1,229 individuals registered in the IBK database from November 8, 2018 through October 17, 2023. Of these, this study cohort considered 419 symptomatic individuals (364 males and 55 females) who harbored a full mutation (355 males and 52 females) or mosaicism (9 males and 3 females), all confirmed by molecular testing based on polymerase chain reaction (PCR) or Southern blotting. For the symptom distribution analysis, only individuals who exhibited at least one observable symptom at the time of data collection were considered.

The inclusion of these individuals aims to describe the frequency of symptoms only among those who present behavioral, cognitive, or physical manifestations compatible with Fragile X Syndrome (FXS). This approach avoids dilution bias that would occur if individuals without any symptoms were included in the denominator, thereby artificially inflating estimates of symptom absence.

### Checklist validation

The study cohort, all confirmed by molecular testing (PCR or Southern blotting), were designated to validate the screening checklist developed in this study for Fragile X Syndrome (FXS), with analysis of the following 12 symptoms: delayed speech; learning difficulties; attention deficit; intellectual disability (ID); hyperactivity; aggressive behavior; avoidance of eye contact; avoidance of physical contact; intentional, repetitive, and rhythmic movements (stereotypies); joint hypermobility; enlarged testes (macroorchidism; considered only for samples composed exclusively of males); elongated face, prominent mandible, and/or protruding ears. All individuals who did not have a confirmed diagnosis, that are premutation carriers, are in the intermediate (gray) zone, or who had a PCR-confirmed negative diagnosis were excluded (negative patients were considered for control purposes). Ethnicity and age were not considered in this analysis.

Data were collected over five years (2018–2023) using an enrollment form that captured personal information, medical history, and the 12 symptoms listed above, which were completed by each individual’s caregiver.

### Study Limitations

This study has limitations stemming from the fact that the registration form was completed by a legal guardian of the individual with FXS rather than by a health professional, which may have introduced variability in the responses. Moreover, data were collected at the time of registration; consequently, individuals’ symptoms may have changed over the ensuing years due to age-related factors.

### Confidence interval

To assess which symptoms are more prevalent among individuals affected by Fragile X Syndrome and which differences between groups are statistically relevant, we computed proportion and 95% confidence intervals (95% CI). For each symptom and group (with or without the syndrome), the proportion was calculated to determine the proportion of individuals presenting the symptom. Subsequently, the standard error of the mean was calculated to quantify the uncertainty of the estimate. To determine the 95% CI, we identified the range within which the true population mean is likely to lie for each group.

Complementary to the 95% CI, to calculate statistical significance (p-value), we compared the prevalence of each symptom between the Positive group (FXS, full mutation) and the Negative group (without Fragile X Syndrome). For this comparison, we applied Pearson’s chi-square test when all expected cell frequencies were ≥ 5, and Fisher’s exact test when any expected frequency was < 5. This approach is appropriate for binary categorical variables (presence/absence of symptoms). Statistical significance was defined as p-value < 0.05.

### Statistical analysis for correlation of signs and symptoms

For the correlation analysis of symptoms among individuals with Fragile X Syndrome, we used Pearson’s correlation coefficient. In this analysis, the closer the correlation coefficient is to +1, the stronger the positive correlation, indicating that if one symptom is present, there is a high probability that another symptom is also present in the same individual. Conversely, the closer it is to −1, the stronger the negative correlation, indicating that the presence of one symptom suggests the other symptom will likely be absent. Finally, if the correlation coefficient is close to 0, this indicates no meaningful relationship between the symptoms.

To visualize the correlations, we generated heatmaps, which provide a visual representation of the correlation matrix across pairs of symptoms (positive or negative correlations).

### Data clustering to identify representative variables in Fragile X Syndrome

For the clustering analysis of symptoms, the dataset was divided into two groups: Positive (FXS, with a full mutation) and Negative (without Fragile X Syndrome). The target variable (group) was then encoded as 1 (Positive) or 0 (Negative). Next, the random forest method (Breiman, 2001) was applied to identify the most important features (symptoms) for distinguishing the two groups. In this context, the top 10 features were selected to reduce dimensionality. For clustering, the k-means method (Rousseeuw, 1987) was used to create two clusters (with and without the syndrome). For visualization, principal component analysis (PCA; Abdi & Williams, 2010) was used to reduce the data to two dimensions, enabling display in a 2D plot.

### Checklist construction criteria

To identify the symptoms that best discriminate the two groups (Positive and Negative) based on statistical significance, we applied statistical analysis and machine learning methods to the data from individuals belonging to both groups. In this context, the collected data were first preprocessed and comprised a set of 620 individuals (201 FXS-negatives; 419 FXS-positives), each characterized by the 12 FXS-associated symptoms.

Next, individuals were divided into two main groups based on condition: Positive Group (symptomatic individuals with full mutation, including mosaic cases) and Negative Group (individuals without Fragile X Syndrome). In addition, the data were stratified by biological sex (male, female) to enable separate analyses, given that symptom manifestation may differ by sex.

Subsequently, the relative frequency of each symptom was computed in the Positive and Negative groups. The relative frequency of a symptom in the Fragile X group (FX) was defined as: FX = N_symptom_FXS / N_FXS, where N_symptom_FXS is the number of individuals in the Positive group who present the symptom, and N_FXS is the total number of individuals in the Positive group.

Analogously, the relative frequency in the Negative group (FN) was calculated as: FN = N_symptom_NFXS / N_NFXS, where N_symptom_NFXS is the number of individuals in the Negative group with the symptom, and N_NFXS is the total number of individuals in the Negative group. The weight of each symptom (P) was then determined as the difference between the relative frequencies in the two groups: P = FX − FN. Symptoms with positive weights are more associated with Fragile X Syndrome, whereas symptoms with negative weights are more associated with the absence of the syndrome.

For each individual, a score was calculated as the sum of the weights of the symptoms present. Formally, the score (S) for an individual i was defined as: , where P_j_ is the weight of symptom j, X_ij_ equals 1 if symptom j is present in individual i and 0 otherwise, and n is the total number of symptoms.

### Determination of the classification threshold for checklist application

To determine the score threshold that classifies an individual as likely affected by Fragile X Syndrome, we used ROC (Receiver Operating Characteristic) curve analysis (Zweig & Campbell, 1993; Fawcett, 2006). The ROC curve was constructed by plotting the true positive rate (sensitivity) against the false positive rate (1 − specificity) for different score threshold values.

The threshold was set to achieve 95% sensitivity, ensuring that 95% of FXS cases were correctly classified, while maintaining specificity. The threshold value (T) was determined as: 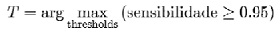

Given T, individuals were classified as Positive for Fragile X Syndrome if their score was greater than or equal to the threshold (S_i_≥T), or Negative for Fragile X Syndrome if their score was below the threshold (S_i_ <T).

To evaluate the method, we assessed the quality of the classification model using the area under the ROC curve (AUC). The AUC ranges from 0.5 (random classification) to 1.0 (perfect classification), with higher values indicating better model performance. In addition, classification performance was assessed by counting true positives (individuals correctly classified as Positive) and false positives (individuals incorrectly classified as Positive).

## RESULTS AND DISCUSSION

### Frequency distribution of the most prevalent symptoms in Fragile X Syndrome

Based on the analysis of signs and symptoms from 419 symptomatic individuals with Fragile X Syndrome, 12 symptoms were evaluated (Table 1, Figures 2–3): learning difficulties (86.87%), attention deficit (79.71%), delayed speech (73.27%), intellectual disability (ID) (72.79%), and intentional, repetitive, and rhythmic movements (70.17%). In descending order of incidence, we then observed elongated face, prominent mandible, and/or protruding ears (65.39%); hyperactivity (59.19%); avoidance of eye contact (52.27%); joint hypermobility (hyperextensibility) (42.24%); avoidance of physical contact (34.61%); and aggressive behavior (31.50%).

**Figure 2.**
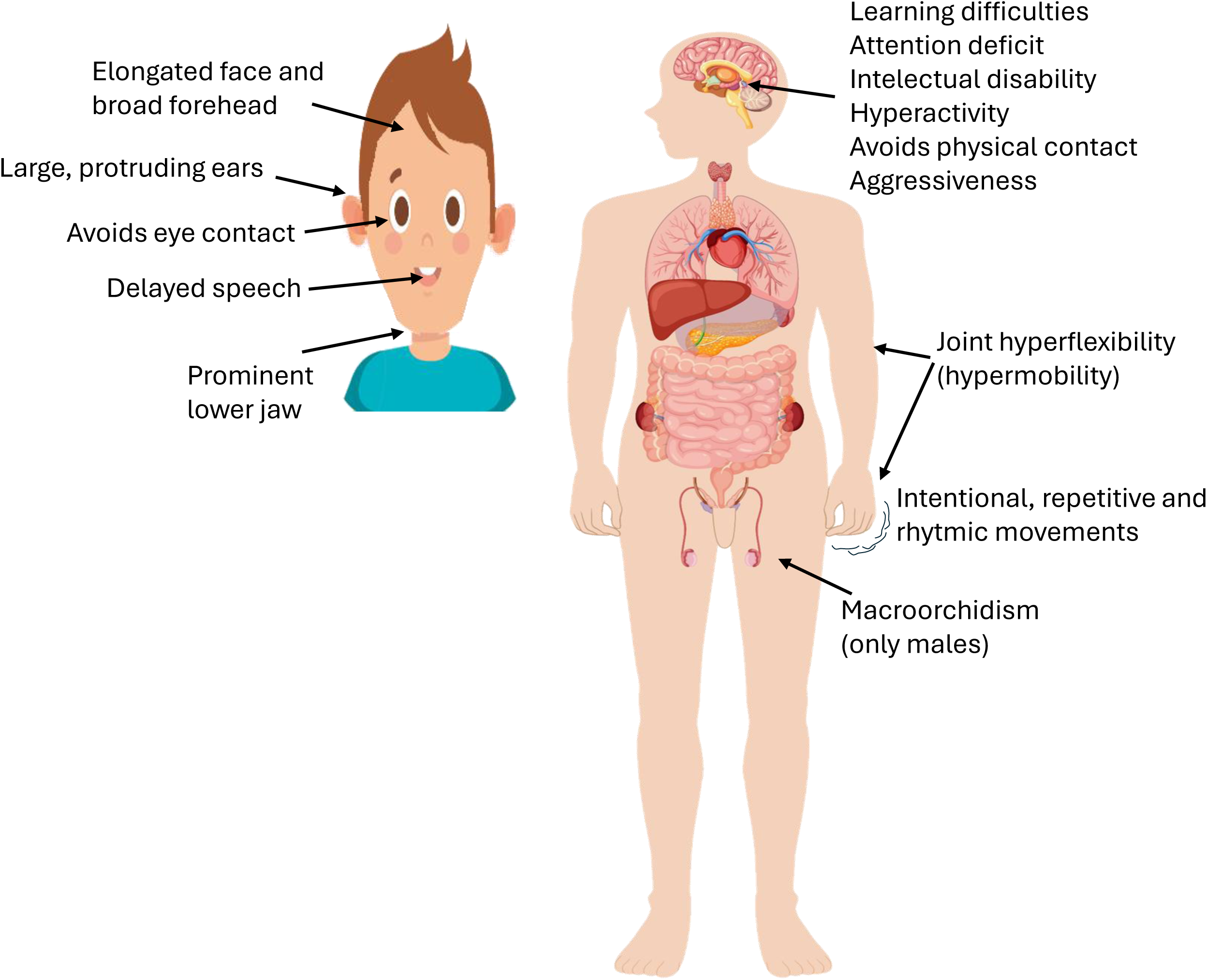
Signs and symptoms of Fragile X syndrome full-mutated individuals (males and females).

**Figure 3.**
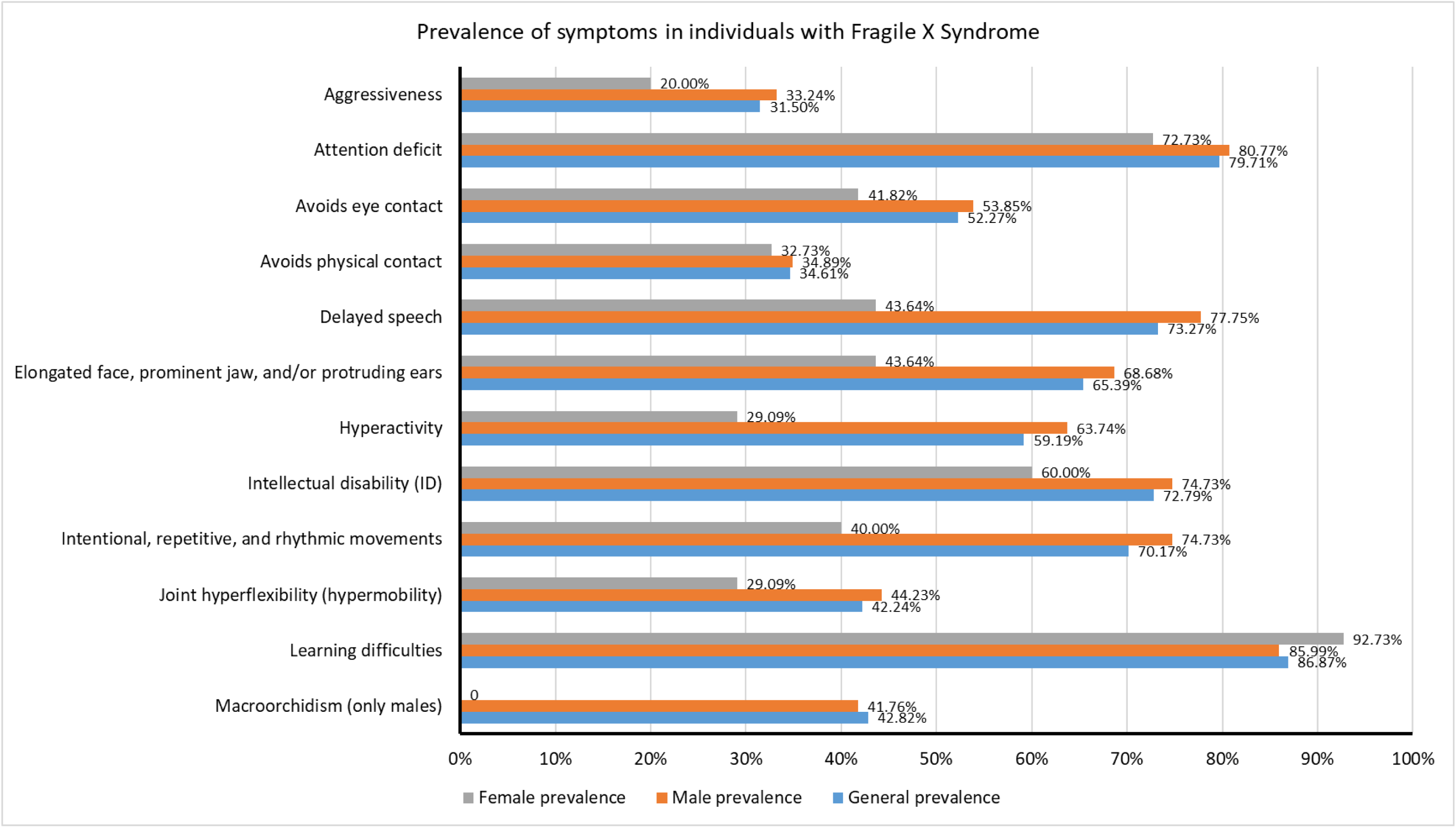
Overall prevalence of clinical characteristics among symptomatic individuals with Fragile X Syndrome (FXS) in Brazil.

**Table 1.** Overall prevalence of clinical characteristics in the male population with Fragile X Syndrome (FXS) in Brazil.

| Male prevalence |  |  |
| --- | --- | --- |
| Learning difficulties | 85.99% | 313/364 |
| Attention deficit | 80.77% | 294/364 |
| Delayed speech | 77.75% | 283/364 |
| Intellectual disability (ID) | 74.73% | 272/364 |
| Intentional, repetitive, and rhythmic movements | 74.73% | 272/364 |
| Elongated face, prominent jaw, and/or protruding ears | 68.68% | 250/364 |
| Hyperactivity | 63.74% | 232/364 |
| Avoids eye contact | 53.85% | 196/364 |
| Joint hyperflexibility (hypermobility) | 44.23% | 161/364 |
| Macroorchidism (only males) | 41.76% | 152/364 |
| Avoids physical contact | 34.89% | 127/364 |
| Aggressiveness | 33.24% | 121/364 |

**Table 2.** Overall prevalence of clinical characteristics in the female population with Fragile X Syndrome (FXS) in Brazil.

| Female prevalence |  |  |
| --- | --- | --- |
| Learning difficulties | 92.73% | 51/55 |
| Attention deficit | 72.73% | 40/55 |
| Intellectual disability (ID) | 60.00% | 33/55 |
| Delayed speech | 43.64% | 24/55 |
| Elongated face, prominent jaw, and/or protruding ears | 43.64% | 24/55 |
| Avoids eye contact | 41.82% | 23/55 |
| Intentional, repetitive, and rhythmic movements | 40.00% | 22/55 |
| Avoids physical contact | 32.73% | 18/55 |
| Hyperactivity | 29.09% | 16/55 |
| Joint hyperflexibility (hypermobility) | 29.09% | 16/55 |
| Aggressiveness | 20.00% | 11/55 |

In the analysis of the sample of 364 male individuals with Fragile X Syndrome, no major divergence was observed in the predominant signs compared with the 419 individuals in the overall cohort. Among these men, the predominant signs were: learning difficulties (85.99%), attention deficit (80.77%), delayed speech (77.75%), intellectual disability (ID) (74.73%), intentional, repetitive, and rhythmic movements (stereotypies) (74.73%), elongated face, prominent mandible, and/or protruding ears (68.68%), hyperactivity (63.74%), avoidance of eye contact (53.85%), joint hypermobility (hyperextensibility) (44.23%), enlarged testes (macroorchidism) (41.76%), avoidance of physical contact (34.89%), and aggressive behavior (33.24%).

Among the 55 female individuals, the distribution of signs diverged from the overall and male prevalence, with the predominant characteristics being: learning difficulties (92.73%), attention deficit (72.73%), intellectual disability (ID) (60%), delayed speech (43.64%), elongated face, prominent mandible, and/or protruding ears (43.64%), avoidance of eye contact (41.82%), intentional, repetitive, and rhythmic movements (stereotypies) (40%), avoidance of physical contact (32.73%), hyperactivity (29.09%), joint hypermobility (hyperextensibility) (29.09%), and aggressive behavior (20%).

For control purposes, we also compared data from the 419 individuals in the overall cohort with Fragile X Syndrome (full mutation confirmed by genetic testing—positive sample) with data from 201 individuals without FXS (no full mutation and diagnosed with Autism Spectrum Disorder (ASD) or another neurological condition distinct from FXS). This comparison indicated that most characteristics—9 of the 12 evaluated—were more prevalent in the FXS-positive sample (Figure 4).

**Figure 4.**
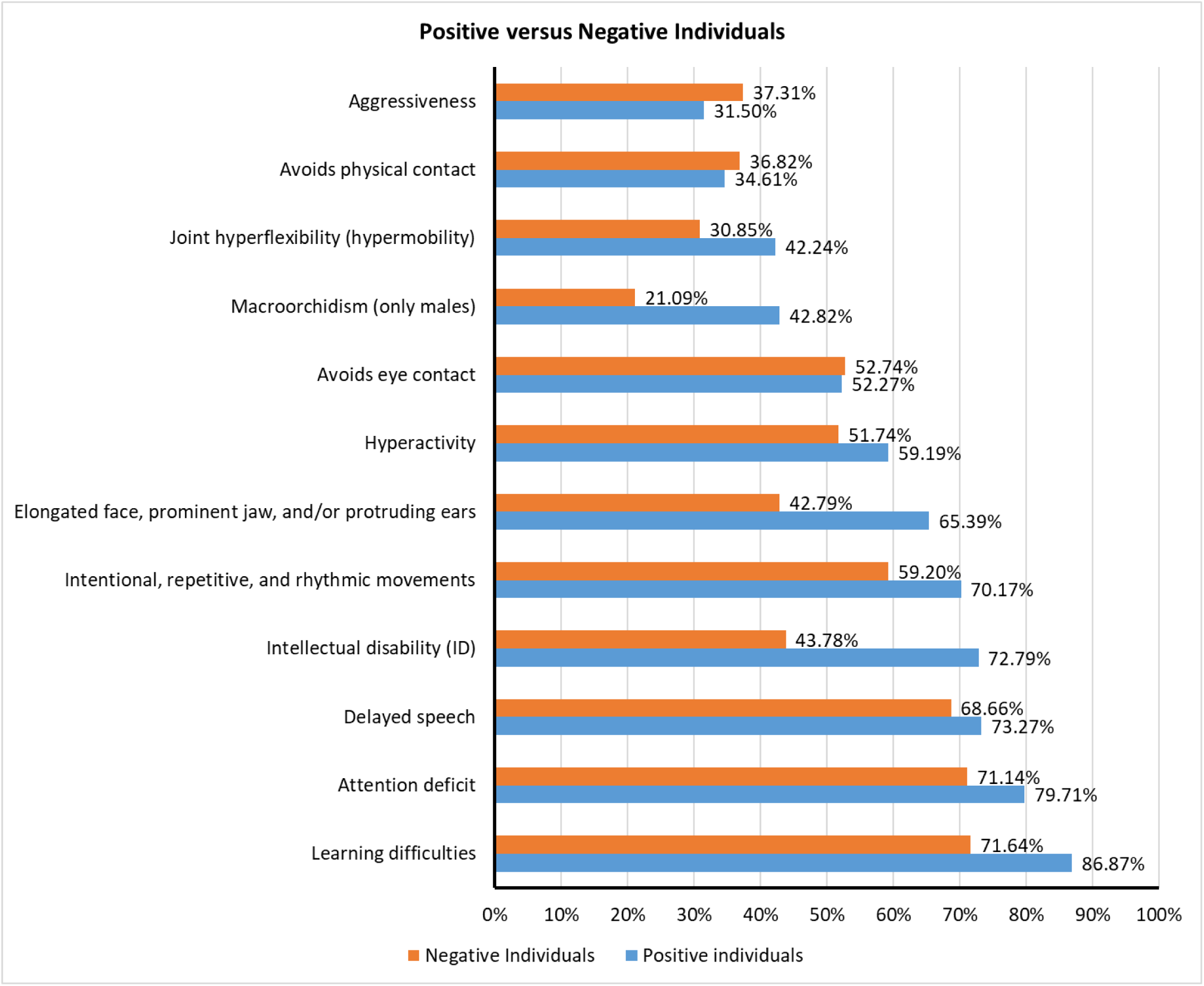
Comparison of checklist characteristics between the Fragile X Syndrome (FXS) group and the FXS-negative group.

### Determination of symptoms with significant occurrence in FXS

Based on the individual-level data, we also conducted an analysis comparing positive cases of Fragile X Syndrome with negative cases (Table 3, Figure 5), reporting 95% confidence intervals (95% CI) for symptom occurrence. For the symptom “attention deficit,” the Positive group shows a mean of 0.80 (i.e., 80% of individuals in this group exhibit the symptom). In contrast, the Negative group has a significantly lower mean, around 0.64 (i.e., 64% of individuals in this group exhibit the symptoms). The error bars do not overlap, indicating a statistically significant difference between groups (p-value < 0.001).

**Figure 5.**
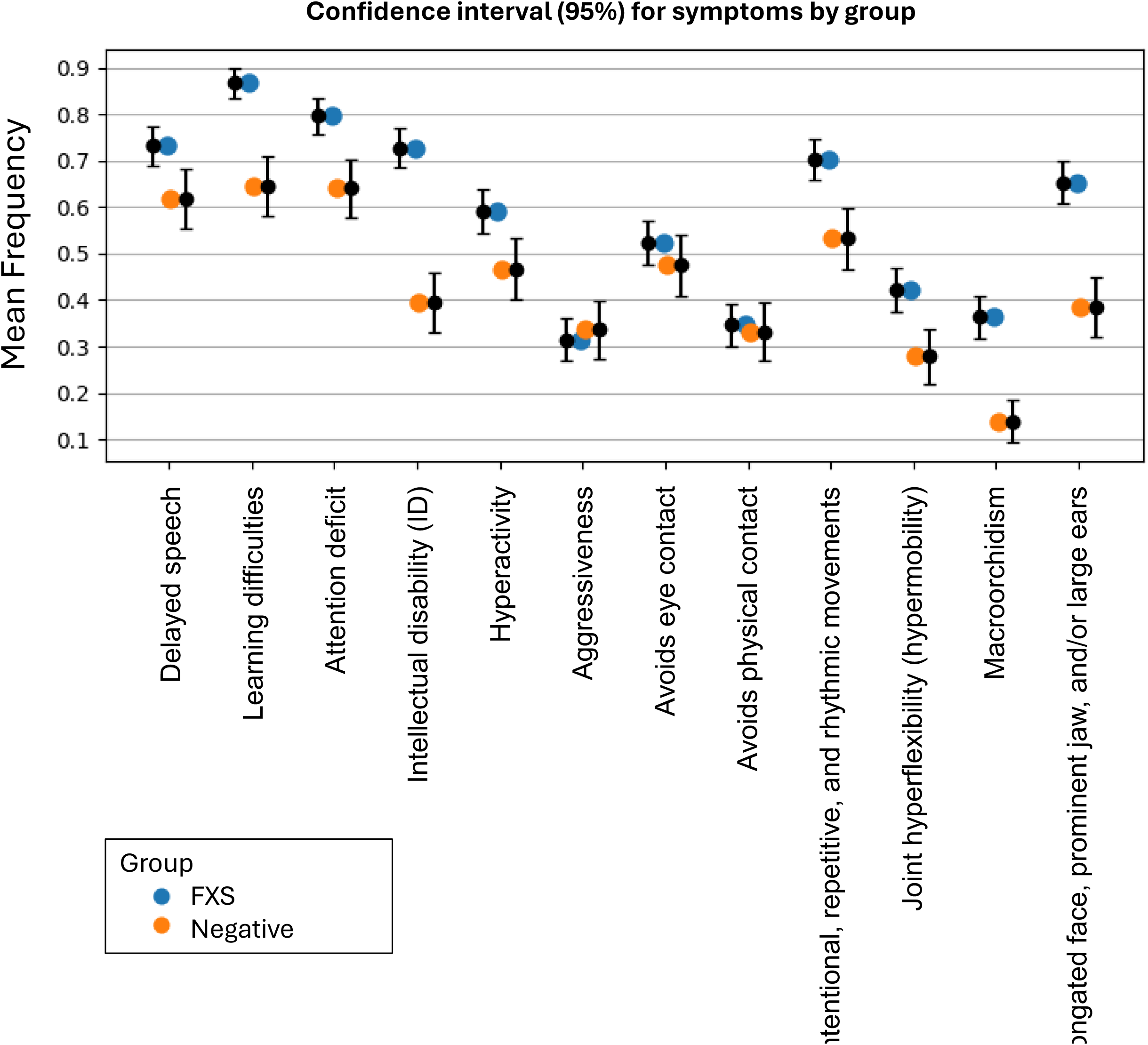
Confidence intervals for symptoms in individuals from the Positive (full mutation) and Negative (no full mutation) groups. For each symptom, the occurrences in the Positive group (blue) and Negative group (orange) are shown. The vertical bar next to each group indicates the mean frequency.

**Table 3.** Confidence intervals for symptoms between the FXS-positive and FXS-negative groups.

| Group | FXS Group |  |  |  | Negative Group |  |  |  | Significance |
| --- | --- | --- | --- | --- | --- | --- | --- | --- | --- |
| Symptom | Mean | Lower CI | Upper CI | Error | Mean | Lower CI | Upper CI | Error | p-value |
| Delayed speech | 0.73 | 0.69 | 0.78 | 0.04 | 0.62 | 0.55 | 0.68 | 0.06 | 0.0039 |
| Learning difficulties | 0.87 | 0.84 | 0.90 | 0.03 | 0.65 | 0.58 | 0.71 | 0.06 | 0.0000 |
| Attention deficit | 0.80 | 0.76 | 0.84 | 0.04 | 0.64 | 0.58 | 0.70 | 0.06 | 0.0000 |
| Intellectual disability (ID) | 0.73 | 0.69 | 0.77 | 0.04 | 0.39 | 0.33 | 0.46 | 0.06 | 0.0000 |
| Hyperactivity | 0.59 | 0.54 | 0.64 | 0.05 | 0.47 | 0.40 | 0.53 | 0.07 | 0.0031 |
| Aggressiveness | 0.32 | 0.27 | 0.36 | 0.04 | 0.34 | 0.27 | 0.40 | 0.06 | 0.6450 |
| Avoids eye contact | 0.52 | 0.47 | 0.57 | 0.05 | 0.48 | 0.41 | 0.54 | 0.07 | 0.2894 |
| Avoids physical contact | 0.35 | 0.30 | 0.39 | 0.05 | 0.33 | 0.27 | 0.39 | 0.06 | 0.7837 |
| Intentional, repetitive, and rhythmic movements | 0.70 | 0.66 | 0.75 | 0.04 | 0.53 | 0.47 | 0.60 | 0.07 | 0.0000 |
| Joint hyperflexibility (hypermobility) | 0.42 | 0.37 | 0.47 | 0.05 | 0.28 | 0.22 | 0.34 | 0.06 | 0.0004 |
| Macroorchidism | 0.36 | 0.32 | 0.41 | 0.05 | 0.14 | 0.09 | 0.18 | 0.05 | 0.0000 |
| Elongated face, prominent jaw, and/or large ears | 0.65 | 0.61 | 0.70 | 0.05 | 0.39 | 0.32 | 0.45 | 0.06 | 0.0000 |

For hyperactivity, the Positive group has a mean of 0.59 (59%), whereas the Negative group has 0.47 (47%), with a statistically significant difference between the two groups (p-value < 0.001). Another discriminating symptom is “joint hypermobility (hyperextensibility),” with a mean frequency of 0.42 (42%) in the Positive group versus 0.28 (28%) in the Negative group, also statistically significant (p-value < 0.001). Thus, this analysis identified that cognitive and behavioral symptoms—such as attention deficit and hyperactivity—differ significantly between groups. In addition, physical features also show large, statistically significant variation (p-value < 0.05) between the Positive and Negative groups, such as elongated face, prominent mandible, macroorchidism, and hyperextensibility.

### Correlation of symptom occurrence in FXS

In the correlation analysis of symptoms in Fragile X Syndrome (FXS), application of Pearson’s correlation identified symptoms that exhibit positive or negative correlations (Figure 6, Table 4).

**Figure 6.**
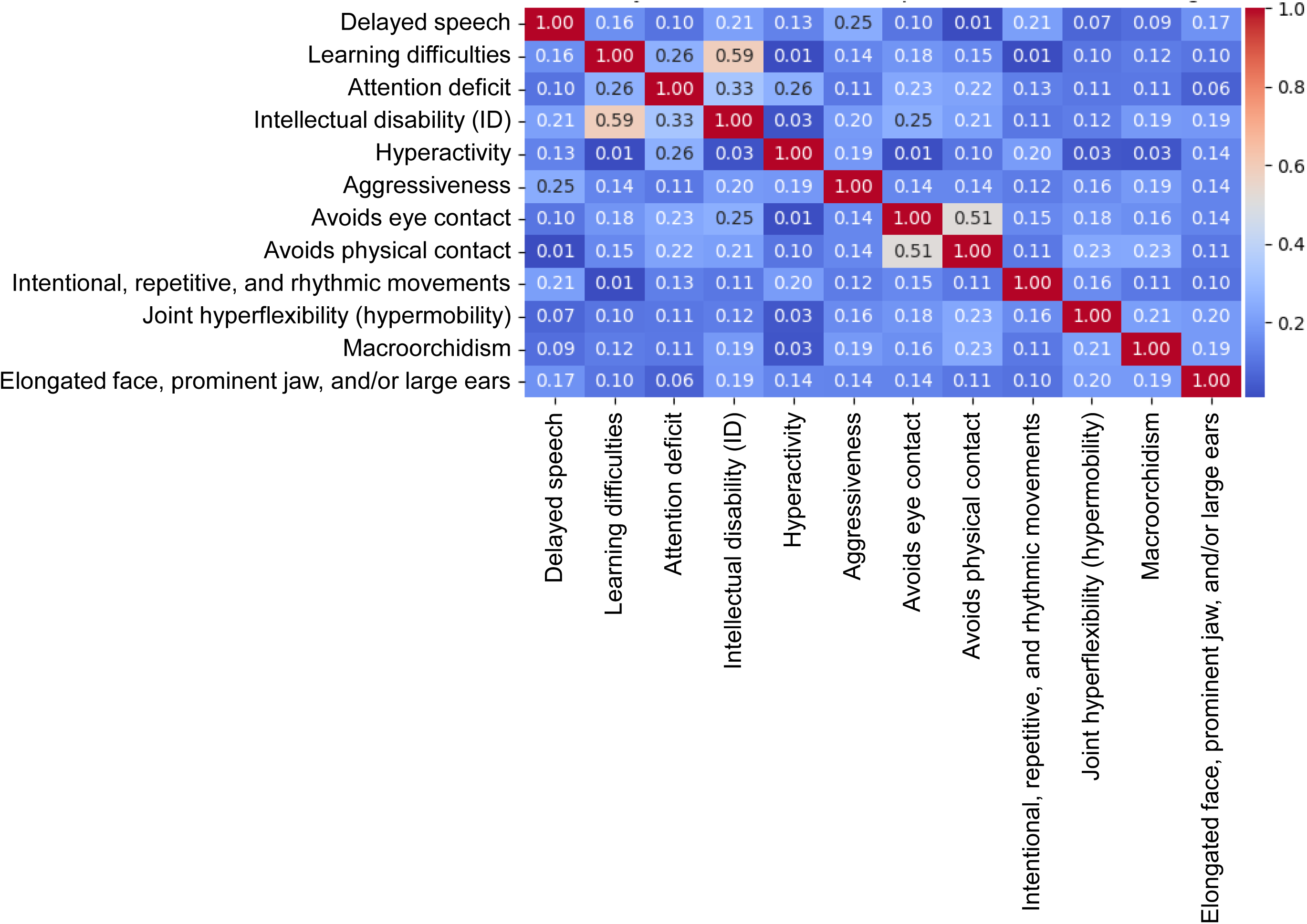
Symptom correlations in Fragile X Syndrome (FXS).

**Table 4.** Symptom checklist with symptom weights for calculating the score of a male individual suspected of having Fragile X Syndrome (FXS).

| Male Symptom Checklist |  |
| --- | --- |
| Symptom | Weight |
| Delayed speech | 0.14 |
| Learning difficulties | 0.18 |
| Attention deficit | 0.17 |
| Intellectual disability (ID) | 0.32 |
| Hyperactivity | 0.12 |
| Aggressiveness | 0.01 |
| Avoids eye contact | 0.06 |
| Avoids physical contact | 0.04 |
| Intentional, repetitive, and rhythmic movements | 0.17 |
| Joint hyperflexibility (hypermobility) | 0.19 |
| Macroorchidism | 0.26 |
| Elongated face, prominent jaw, and/or protruding ears | 0.29 |

Among the correlations identified between pairs of symptoms in individuals with Fragile X Syndrome (FXS), the strongest was between intellectual disability (ID) and learning difficulties, with a moderate positive correlation (r = 0.59). Another pair with a moderate positive correlation was avoidance of eye contact and avoidance of physical contact (r = 0.51). Conversely, no symptom pairs showed moderate or strong negative correlations.

Beyond prevalence analysis, we also investigated co-occurrence patterns of clinical manifestations to better understand how symptoms cluster in individuals with Fragile X Syndrome. Network-based analysis revealed that cognitive and behavioral symptoms tended to cluster together, with learning difficulties, attention deficit, and intellectual disability forming a core subgroup that co-occurred in more than 70% of cases. Physical characteristics, such as elongated face and macroorchidism, were more sporadically associated with behavioral traits, but consistently co-occurred with intellectual disability. These findings support the existence of symptom clusters rather than isolated traits, reinforcing the need for multidimensional assessment when screening for FXS.

### Development of a Brazilian checklist for Fragile X Syndrome

Based on analyses of data from individuals of both sexes with Fragile X Syndrome (FXS), we developed a checklist to determine whether a person suspected of having the syndrome should be referred for molecular testing. For checklist construction, each symptom—considering both males and females with FXS—was analyzed to compute its respective weight in the syndrome.

For male individuals (Table 4), the weighting of each symptom was calibrated using a score threshold of 0.56 that achieved 95% sensitivity (area under the ROC curve [AUC] for males = 0.73). Among the highest-weighted symptoms are intellectual disability (ID) and elongated face, prominent mandible, and/or protruding ears. The score threshold determines whether an individual is classified as suspected of FXS.

For female individuals (Table 5), the weight assigned to each symptom was calculated using a score threshold corresponding to 95% sensitivity (threshold = 0.55, the same as for males), with an area under the ROC curve (AUC) of 0.76 for this group. Among the highest-weighted symptoms are learning difficulties and intellectual disability (ID).

**Table 5.** Symptom checklist with symptom weights for calculating the score of a female individual suspected of having Fragile X Syndrome (FXS).

| **Female Symptom Checklist** |  |
| --- | --- |
| **Symptom** | **Weight** |
| Delayed speech | 0.01 |
| Learning difficulties | 0.28 |
| Attention deficit | 0.12 |
| Intellectual disability (ID) | 0.20 |
| Hyperactivity | 0.04 |
| Aggressiveness | 0.02 |
| Avoids eye contact | 0.08 |
| Avoids physical contact | 0.07 |
| Intentional, repetitive, and rhythmic movements | 0.05 |
| Joint hyperflexibility (hypermobility) | 0.04 |
| Elongated face, prominent jaw, and/or protruding ears | 0.09 |

Based on the presented weights, to classify an individual as “suspected” of having Fragile X Syndrome based on their symptoms, one simply evaluates the symptoms in the table and accumulates and sums the points for all symptoms present. If the score exceeds the threshold value of 0.55, this indicates that the individual is suspected of having Fragile X Syndrome and should undergo genetic testing.

For example, for a male individual who presents the symptoms of intellectual disability (ID), learning difficulties, enlarged testes, hyperactivity, and aggressive behavior, the score is calculated as follows: P = 0.32 + 0.18 + 0.26 + 0.12 + 0.01 = 0.89. Since P = 0.89 is greater than the male threshold of 0.56 (95% sensitivity), this individual is classified as suspected of Fragile X Syndrome.

To illustrate for female individuals, suppose a person presents the symptoms of ID, hyperactivity, and delayed speech; her score will be: P = 0.20 + 0.04 + 0.01 = 0.25. Because P = 0.25 is below the score threshold of 0.55 (95% sensitivity) for females, this person is considered negative for suspected Fragile X Syndrome, and therefore specific genetic testing for the syndrome is not recommended.

## CONCLUSION

This study comprehensively assessed the prevalence of physical signs and clinical symptoms among individuals affected by Fragile X Syndrome (FXS). It encompasses the largest cohort of symptomatic individuals with a full mutation or mosaicism studied worldwide to date (n = 419) and constitutes the largest investigation conducted in the Brazilian population.

Using symptom data from individuals with FXS, we performed a series of statistical analyses and observed several symptoms with prevalence above 70% in FXS, including learning difficulties, attention-deficit symptoms, delayed speech, and intentional, repetitive, and rhythmic movements. We also found that the symptoms with the highest prevalence in males and females with FXS occur at broadly similar frequencies and are the most common in both groups, apart from sex-specific manifestations such as macroorchidism.

In addition, we conducted an extensive analysis of data from individuals with FXS to create a Brazil-based checklist that reflects the characteristics of FXS in a highly admixed and ethnically diverse population. The resulting 12-symptom checklist serves as a practical decision-support tool to guide clinical evaluation and highlight which aspects should prompt a health professional to recommend molecular testing for syndrome detection.

Although the checklist covers a limited set of symptoms, integration with additional clinical and family history information is essential, such as a family history of intellectual disability, the occurrence of autism spectrum disorder, epilepsy, infertility, premature menopause, and recurrent miscarriage. These additional factors may signal the possibility of genetic inheritance—whether due to a premutation or a full mutation in the FMR1 gene—directly associated with Fragile X Syndrome.

Finally, this study differs from prior work by analyzing the frequency of signs and symptoms in individuals with a confirmed diagnosis of FXS (full mutation or mosaicism, both established molecularly). Unlike most earlier studies that applied pre-existing checklists before conducting molecular testing of suspected cases, our investigation began with individuals already confirmed to have FXS (full mutation or mosaicism, defined molecularly). In this context, the present work confers direct relevance to the symptoms identified in individuals with FXS and delineates clinical “red flags” that should trigger molecular testing, while accounting for the syndrome’s generalized phenotype and adapting to the Brazilian setting.

## Data Availability

All data produced in the present work are contained in the manuscript.

## ACKNOWLEDGEMENTS

RHH is supported by Fundação Araucária (FA, grant #CP09/2016) and Conselho Nacional de Desenvolvimento Científico e Tecnológico (CNPQ, grant #311438/2022-9 and #402773/2022-5).

## ETHICAL APPROVAL

This research was approved by the Ministry of Health of Brazil through the Human Research Ethics Committee (CEP) (CAAE: 92396125.3.0000.0020).

